# Relation of frailty with depression among Colombian COPD adults aged over 60 years

**DOI:** 10.1101/2024.02.29.24303577

**Authors:** Adalberto Campo-Arias, John Carlos Pedrozo-Pupo, Carmen Cecilia Caballero-Domínguez

## Abstract

**Introduction:** Frailty and depression risk are common in older adults undergoing chronic obstructive pulmonary disease (COPD) treatment. However, little is known about this association in people with COPD residing in low- and middle-income countries. The study aimed to o establish the relationship between frailty and depression among ambulatory adults over 60 years with COPD in Santa Marta, Colombia.

**Methods:** A cross-sectional study was designed in which consecutive patients from the pulmonology outpatient clinic were invited to participate. Frailty was quantified with the FiND (Frail Non-Disabled [FiND] Screening Tool) [Cronbach’s alpha 0.65] and depression with the Primary Care Screening Questionnaire for Depression (PSQD) [Cronbach’s alpha 0.73].

**Results:** Three-hundred forty-nine patients with COPD between 60 and 98 years participated (M=75.6, SD=8.4), 61.9% were males, and 19.8% presented a C or D combined evaluation. Two hundred eighty-six patients (76.8%) presented frailty with and without mobility disability, and 31.2% presented depression. The relationship of frailty with depression remained significant, even after adjusting for confounding variables (OR=2.80, 95%CI 1.42-5.51).

**Conclusions:** Frailty and depression are significantly associated after adjusting for confounding variables. More studies are welcome.

## 1 INTRODUCTION

Frailty affects up to 80%, quantified with the Frail Non-Disabled (FiND) Screening Tool, of people living with chronic obstructive pulmonary disease (COPD). Patients living with COPD are twice as likely to be frail.^1^ The prevalence of depression is widely variable among people with COPD; it can be identified in up to 60 % of patients.^2-4^ Depression is three to four times more likely among COPD patients than the general population.^5^ Depression is nearly 40% among frail people;^6^ these patients have seven times the depression risk.^7-9^

Frailty and depression are reciprocal relationship^10^ or can be part of a single syndrome.^11^ Frailty, depression, and COPD interact synergistically to increase prolonged hospital stays and suicidal behavior.^12^ Then, a comprehensive approach to patients with comorbidity favors treatment adherence increases social adaptation, and improves patients’ quality of life.^13^

The relationship between frailty and depression has various studies with a positive association among community-dwelling adults.^9^ However, the association between frailty and depression in patients with COPD has been little studied worldwide. It is necessary to consider these entities together to understand better the association’s complexity and the specific clinical needs of this group of patients.^13^ This comorbidity substantially increases the deterioration in the quality of life and the risk of hospitalization and death.^11^

With the participation of Colombian patients, this study can strengthen the knowledge of the association between frailty and depression if cultural particularities are considered. Cultural factors may favor coping with illness and the risk of depression. Colombia has many extended families with several generations sharing the same house with a high frequency of familism. It is a cultural value emphasizing support, attachment, and loyalty to the family.^14^

The study aimed to determine the relationship between frailty and depression among ambulatory adults over 60 years with COPD from Santa Marta, Colombia.

## 2 METHODS

### 2.1 Design and ethical issues

A cross-sectional study was designed. The Research Ethics Committee of the Universidad del Magdalena at Santa Marta, Colombia, approved the study protocol according to minutes 003 of an ordinary virtual session held on March 4, 2021. All patients signed written informed.

### 2.2 Participants

Adult patients older than 60 years with a diagnosis of COPD were included. Patients with evident cognitive impairment and participants with limitations in filling out the self-administered questionnaire for depressive symptoms were excluded.

There was a non-probabilistic sample of patients who consulted between July 1, 2021, and June 30, 2022. Demographic variables, COPD combined evaluation, depression, and frailty were measured. The four stages of combined evaluation were classified into two groups: the less severe (A and B) and the more severe patients (C and D).^15^

### 2.3 Measures

Depression was quantified with the Primary Care Screening Questionnaire for Depression (PSQ4D). The PSQ4D self-report tool includes four items that explore mood during the two most recent weeks. This instrument has a dichotomous response pattern; each affirmative response receives one point. Scores of two or more indicate a risk of depression with a specificity of 0.96 and sensitivity of 0.87.^16^ The PSQ4D presented a Cronbach’s alpha of 0.73 in the present sample.

Frailty was measured with the FiND, which includes five items, the first two for ‘disability’ and the remaining three for ‘frailty.’ Each item has two scoring options, zero and one. If item 1 + item 2 ≥ 1, the individual is considered ‘disabled.’ The individual is considered ‘frail’ if item 1 + item 2 = 0 and item 3 + item 4 + item ≥ 1. The individual is considered ‘strong’ if the sum of all items is zero. The FiND presents a specificity of 0.76 for identifying non-disabled and disabled frail participants.^17^ Frail patients with and without mobility disability were pooled and compared to non-frail patients. The FiND showed a Cronbach’s alpha of 0.65 in the present study.

### 2.4 Data analysis

Frailty was used as the independent variable, and depression as the dependent variable. Demographic and combined assessment variables were taken as confounding variables. Odds ratio (OR) with 95% confidence intervals (95%CI) were calculated. Adjustment for the association between frailty and depression was performed using logistic regression. The analysis was performed in the IBM SPSS Statistics for Windows, Version 23.0.^18^

## 3 RESULTS

Three hundred and forty-nine patients aged between 60 and 98 years (M=75.6, SD=8.4) participated. See Table 1. The disability scores were between 0 and 2 (M=0.74, SD=0.87), and 163 (46.7%) patients were classified as people with disability. Frailty scores were observed between 0 and 3; 105 (30.1%) presented frailty. Consequently, 286 patients (76.8%) were grouped as frail patients with and without mobility disability. The depression scores were between 0 and 4 (M=1.1, SD=1.3), and 109 patients (31.2%) presented depression.

**Table 1.** Demographical and clinical features of the sample.

| Variable | n | % |
| --- | --- | --- |
| Age between 60 and 79 years | 227 | 65.0 |
| Female gender | 133 | 38.1 |
| High school or less | 272 | 77.9 |
| Married or free union | 309 | 88.5 |
| Low income | 258 | 73.9 |
| Significant deterioration (C or D combined evaluation) | 69 | 19.8 |

The association between frailty and depression was statistically significant (OR=3.26, 95%CI 1.68-6.32). However, the gender and combined evaluation behaved as confounding variables. The relationship between frailty and depression remained significant, even after adjusting for confounding variables (OR=2.80, 95%CI 1.42-5.51).

## 4 DISCUSSION

The present significant relationship between frailty and depression among Colombia COPD patients is similar to those previously observed. From a meta-analysis, Vaughan et al.^11^ concluded that patients with frailty had almost three times the risk of depression.

Frailty may represent a significant stressor leading to depression during the illness.^11^ Decreased physical activity during depression may represent a risk for frailty.^10^ Finally, frailty and depression could be manifestations of a single syndrome.^13^ Further studies should always control for confounding variables.

The comorbidity of frailty and depression among COPD patients worsens lung disease, impairs quality of life, and increases mortality. Particular attention should be paid to depression due to its close relationship with suicidal behaviors.^12,19^ The timely identification and treatment of comorbidity significantly improve the quality of life and long-term prognosis of people with COPD.^13^

The present study has the novelty of corroborating the association between frailty and depression in a sample of middle-income countries in South America with prevalent familism that could affect the positive association between frailty and depression documented in high-income countries.^14^ However, it has some limitations: cross-sectional design, non-probabilistic sample, and non-formal depression diagnosis.^20^

In conclusion, frailty triples the likelihood of depression among people with COPD. There is a need to explore frailty and depression among COPD patients systematically. Longitudinal studies can clarify the causal relationship.

## Data Availability

Data supporting the findings of this study are available upon reasonable request to the corresponding author.

